# Quantitative 4D-Flow MR Imaging of Abdominopelvic Vasculature in Pelvic Venous Disorders

**DOI:** 10.1101/2025.08.20.25334119

**Authors:** Christine E. Boone, Sophie Wong, Pam Taub, Anne C. Roberts, Christopher R. Ingraham, Albert Hsiao, Joy Liau

## Abstract

**Purpose:** To determine whether quantitative 4-Dimensional (4D)-Flow MRI could reflect morphologic findings of pelvic venous disorder (PeVD).

**Methods:** Abdominopelvic MRI with 4D-Flow acquired with 3T MRI from 2016-2022 were retrospectively reviewed for morphologic imaging findings: no venous abnormalities (NVA), left common iliac vein compression, left gonadal vein reflux, left renal vein (LRV) compression, and presence of pelvic collaterals. Using 4D-Flow MRI, blood flow was measured for vascular segments from the level of the suprarenal inferior vena cava (IVC) to the common iliac veins. Flow measurements at the LCIV and right common iliac vein (RCIV), the perihilar and juxta-caval renal veins were compared among participants with NVA, LCIV compression, LRV compression with and without LGV reflux, and with LGV reflux without LRV compression.

**Results:** Sixty-six participants with LCIV compression, LRV compression, or LGV reflux displayed significantly diminished flow adjacent to the site of compression or reflux. Compared to participants with NVA, those with LCIV compression with pelvic collaterals showed increased RCIV flow and decreased LCIV flow (LCIV:RCIV flow ratio: 0.49±0.08 vs. NVA:0.92±0.05, p=0.0005). LCIV compression without pelvic collaterals did not significantly differ from NVA (LCIV:RCIV flow ratio: 0.80±0.09 vs. NVA, p>0.1). LRV compression with LGV reflux showed diminished juxta-caval LRV flow and similar perihilar LRV flow compared to LRV compression without LGV reflux ( p=0.03) or NVA (p=0.004) (juxta-caval:perihilar LRV flow ratio: with LGV reflux:0.39±0.17, without LGV reflux:1.3±0.19, NVA:1.3±0.13).

**Conclusions:** Quantitative abdominopelvic 4D-Flow MRI measurements reflected flow diversion away from obstruction in LCIV or LRV compression, particularly in the setting of decompressing venous reservoirs.

## Introduction

Pelvic venous disorder (PeVD) is an underrecognized cause of chronic pelvic pain, characterized by pelvic venous stasis and varices. Endovascular treatment of a pelvic venous disorder (PeVD) is patient-specific, with treatment consisting of targeted embolization of dilated veins and sites of venous reflux, and stenting sites of obstruction [1–6]. Causes for treatment failure with residual or worsened symptoms can be multifactorial, but re-canalization of embolized vessels, formation of new collaterals, or inadvertent embolization of decompressing venous reservoirs are among these [1,2,7,8]. Delineating and localizing venous abnormalities, including compression, reflux, and flow diversion, are essential to optimally address hemodynamic pathophysiology.

The gold diagnostic standard, invasive venography (IVG) often augmented with intravascular ultrasound (IVUS), is dependent on the operator and direct visualization of morphologic and hemodynamic abnormalities. Advanced magnetic resonance imaging (MRI), such as Differential Sub-sampling with Cartesian Ordering (DISCO) [9] and three-dimensional time-resolved phase-contrast MR angiography/venography (MRA/MRV) with velocity encoding (4D-Flow MRI), enable acquisition of high spatiotemporal multiphasic contrast-enhanced images providing dynamic visualization of vasculature and quantitative blood flow assessment in vascular abnormalities [10–18], cerebrovascular [19,20], cardiac [21–29], and hepatic and portal venous systems [30–32]. Quantitative flow evaluation may detect difficult to visualize venous collaterals or reflux sites. The conservation of flow principle, also utilized in evaluation of structural cardiovascular abnormalities, can reveal aberrant blood flow patterns in a quantitative flow analysis [33]. This study assessed whether 4D-Flow MRI-derived hemodynamic data could identify and quantify abdominopelvic venous abnormalities venous compression, reflux, and diverted flow.

## Methods

### Participants

With Health Insurance Portability and Accountability Act compliance and Institutional Review Board approval, 78 clinically indicated abdominopelvic 4D-Flow MRI acquisitions performed at our institution between April 2016 and April 2022 were retrospectively analyzed. Indications included PeVD, May-Thurner Syndrome (MTS)—associated with obstruction at the left common iliac vein (LCIV) by the right common iliac artery, Postural Orthostatic Tachycardia Syndrome (POTS), Ehlers-Danlos Syndrome, obstruction at the left renal vein (LRV)—associated with Nutcracker syndrome, primary gonadal vein (GV) reflux related to incompetent venous valves, or concern for non-vascular pathology (e.g. uterine fibroids) [4,34–36]. Exclusion criteria were venous abnormalities related to nonvascular etiology, arterial abnormalities, or prior endovascular therapy. Sixty-six participants met inclusion criteria for further analyses.

### Data Acquisition

#### MRI

3T abdominopelvic MRI (Discovery MR 750, GE Healthcare) was performed with a 32-channel phased-array coil. After administration of either 0.15 mmol/kg gadolbutrol or 0.1-0.15 mmol/kg gadobenate, postcontrast MRI sequences were acquired including DISCO and LAVA-Flex from the kidneys through the femoral heads. 4D-Flow MRI was acquired with retrospective cardiac gating during free breathing as a coronal slab extending through the abdomen and pelvis with arms positioned above the head [37]. Velocity encoding gradient (VENC) were 80, 100, 120, 150, or 200 cm/s. A wider variety of velocity encoding speeds were used early in our experience with 4D Flow prior to May 2021, which was then standardized a 120 cm/s VENC.

#### IVG

IVG (Artis Zeego, Siemens) was performed via femoral venous access in 17 participants by or under direct supervision of an experienced attending interventional radiologist. Examinations included pelvic venography from the inferior vena cava (IVC) confluence, bilateral iliofemoral venography, bilateral renal venography, and gonadal venography (if visualized). The fluoroscopy table was positioned to 15-degree reverse Trendelenburg.

### Data processing

Manual phase error correction and vessel segmentation for flow quantification was performed using dedicated imaging software (Cardio AI, 26.9.1; Tempus Pixel, Chicago IL). Orthogonal ROIs were made at the suprarenal aorta, suprarenal IVC, infrarenal IVC, juxta-caval left and right renal veins (LRV, RRV), perihilar LRV and RRV, IVC confluence of the left and right common iliac veins (LCIV, RCIV), the proximal LCIV and RCIV (**Figure 1A**). Segmentation and flow measurements at the aortic bifurcation and the bilateral juxta-caval renal arteries were performed. If aliasing was observed, a Tempus Pixel anti-aliasing algorithm was applied. Measurements (net flow and peak speed) were collected over the cardiac cycle and binned into 20 (duration of each range 33-45 ms) or 60 (12-19 ms) equivalent periods. Further details of data processing and segmentation are described in the supplementary methods.

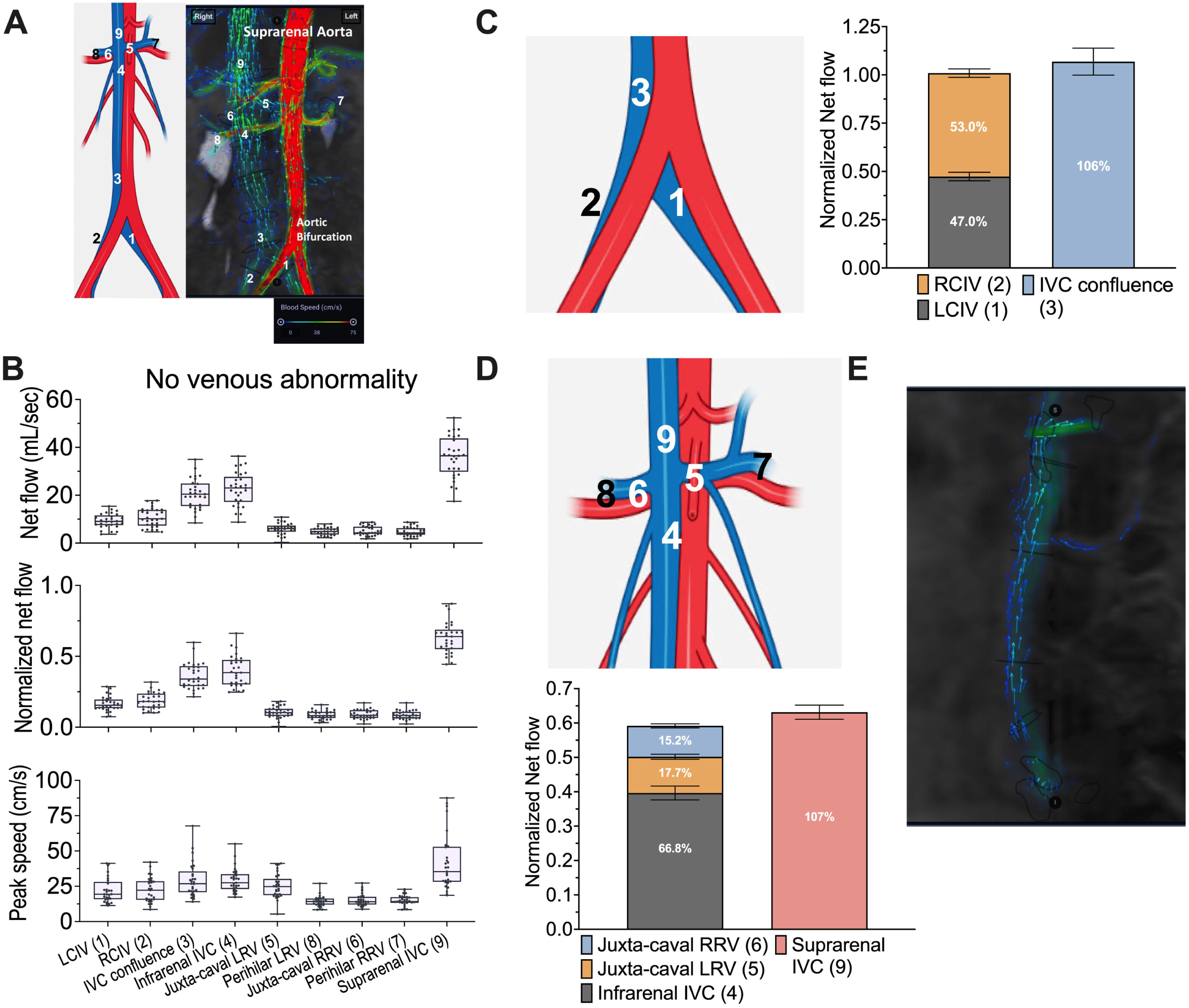

### Classification of participants by morphological imaging findings

Data was extracted from finalized imaging reports from MRI and IVG (when performed). Participants were classified by morphological imaging findings in the reports, which included: normal-appearing vessels/absence of venous abnormalities (NVA), LCIV compression, GV reflux (bilateral, left, or right), and LRV compression. Imaging findings can coexist in the same participants. While these groups were distinct, participants with multiple imaging findings were in multiple groups. Participants were further stratified into LGV reflux without LRV compression, LRV compression without LGV reflux, or LRV compression with LGV reflux groups. LGV reflux participants had either LGV only or bilateral GV reflux. The presence of pelvic collaterals was evaluated. Participants with LCIV compression were stratified into LCIV compression with pelvic collaterals or LCIV compression without pelvic collaterals groups.

### 4D-Flow MRI Quantitative Data Analysis

Data were analyzed in Matlab (Mathworks, R2021a Update 4, 9.10.0.1710957) and GraphPad Prism 10. The mean of net flow and the maximum of peak speed measurements over the cardiac cycle were calculated for each segmented vessel position in each subject. Unless specified otherwise, flow and speed measures are the mean net flow (mL/s) and maximum peak speed (cm/s), respectively. To facilitate comparison between participants, the mean net venous flow was divided by arterial inflow to obtain the normalized net flow (NNF) per participant. Renal venous flow was normalized by the summed mean net flow of the bilateral renal arteries. Common iliac venous flow was normalized to the mean net flow at the aortic bifurcation. Unless specified otherwise, all group comparisons are NNF. The LCIV/RCIV flow ratio is the NNF at the LCIV divided by NNF at the RCIV. The LRV flow ratio is the NNF at the juxta-caval LRV divided by NNF at the perihilar LRV. Further details of quantitative flow analysis are described in the supplementary methods.

## Statistical Analysis

Non-parametric tests were used as some group sizes were too small to visually confirm or test for normality. For comparisons of flow between the NVA group and other groups at different vessel locations, Mann-Whitney test was used with Holm–Sidak’s correction for multiple comparisons. Comparison of one variable across multiple independent groups was performed using Kruskal-Wallis test with Dunn’s multiple comparisons test. Comparison of paired locations across independent groups was performed using Wilcoxon matched pairs test and Holm–Sidak’s correction for multiple comparisons. The significance threshold was set at *p*=0.05. Data are presented as mean ± standard error of the mean (SEM).

## Results

### Participant characteristics and classification

Table I summarizes demographic and imaging findings of 66 participants meeting inclusion criteria for further analyses. No patients had thromboembolic disease.

**Table I.**
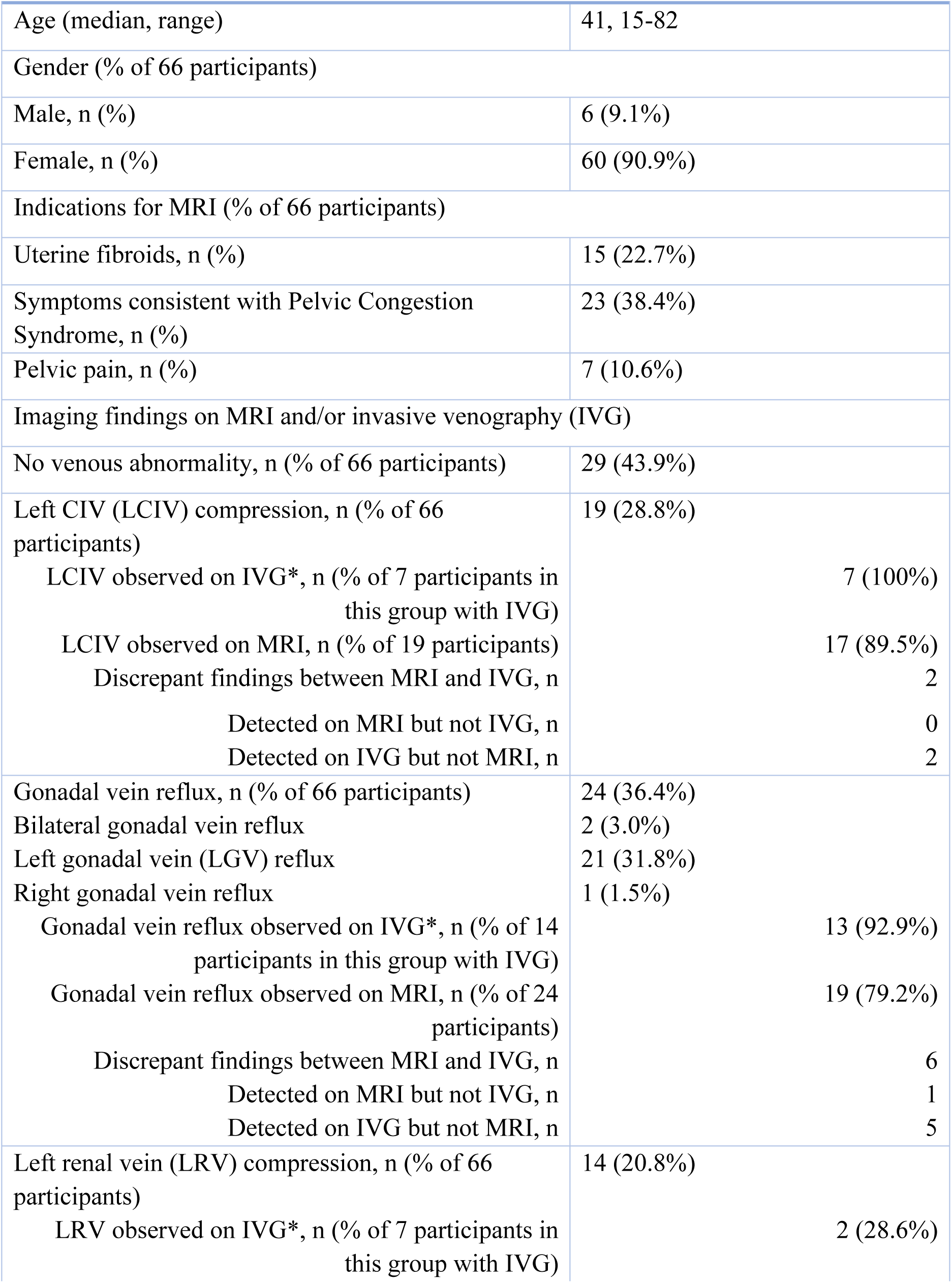
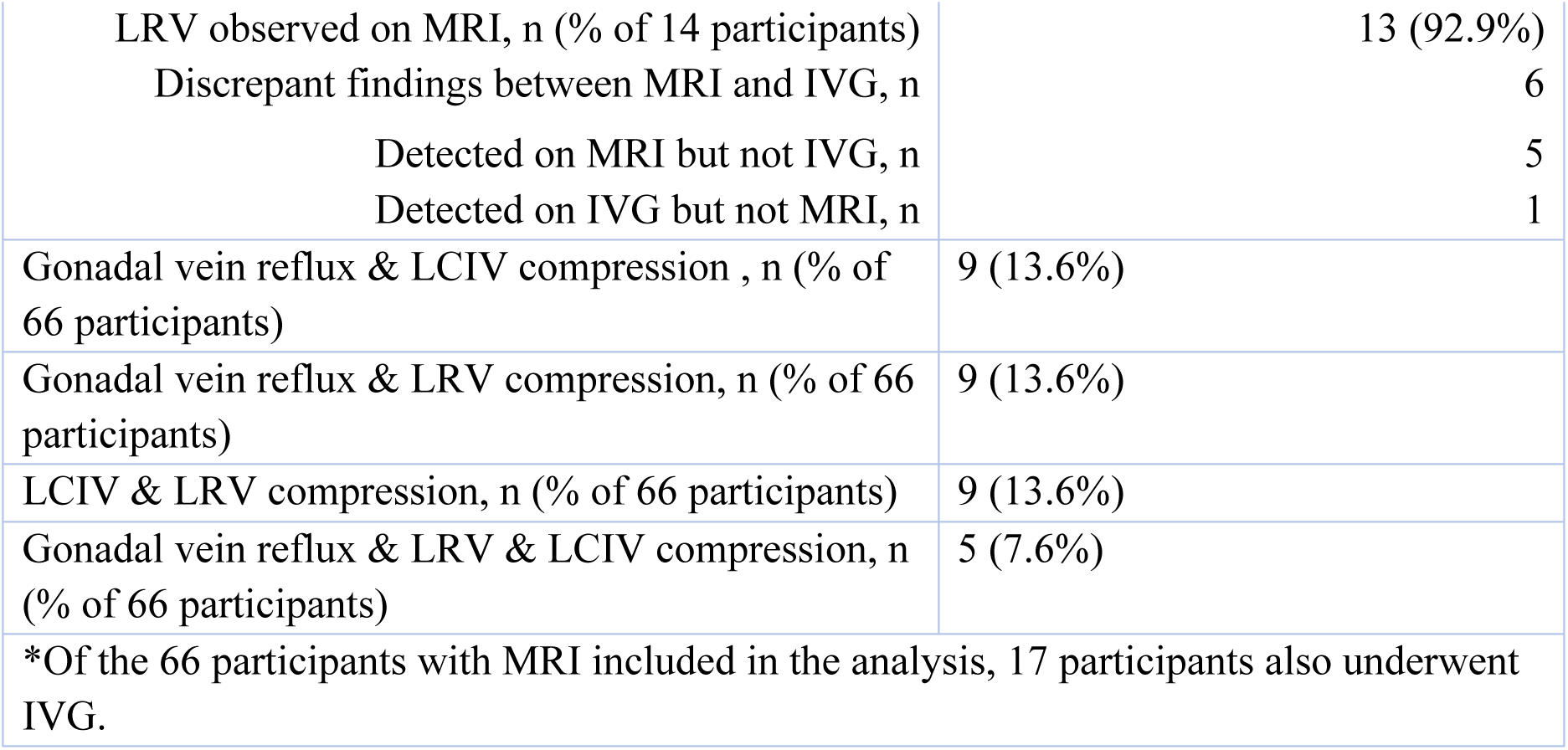
Participant characteristics and Imaging finding classifications.

### Conservation of flow at venous confluences in participants without venous abnormality

Blood flow and peak speed in 29 NVA participants were analyzed at defined locations along the abdominopelvic path of venous return (**Figure 1A**). Non-normalized net flow at the IVC confluence, infrarenal IVC, and suprarenal IVC was 20.6±1.1, 22.6±1.3, and 36.0±1.6 (mean±SEM, mL/s), respectively. Maximum peak speed (cm/s) at the IVC confluence, infrarenal IVC, and suprarenal IVC were 29.3±2.3, 29.2±1.6, and 41.9±3.7, respectively. Non-normalized net flow and peak speed values were concordant with reported MRI and doppler ultrasound measurements.[13,38–40] Net flows normalized to suprarenal aortic net flow showed similar patterns of flow as non-normalized net flow from the CIVs to the suprarenal IVC (**Figure 1B**). NVA participants were grouped by their VENC, and maximum peak speed, non-normalized net flow, and NNF were similar among the groups (**Supplemental Figure 1**).

Conservation of flow was observed. The NNF at venous confluences were similar, but slightly greater than the sum of the NNF measured at the contributing veins. At the IVC confluence, the NNF (1.1±0.07) was slightly greater (6%) than the combined NNF from the LCIV (0.47±0.02) and RCIV (0.54±0.02) (**Figure 1C**). NNF just above the renal vein-IVC confluence (0.63±0.02) was slightly greater (7%) than the combined NNF from the juxta-caval LRV (0.11±0.01), juxta-caval RRV (0.09±0.01), and infrarenal IVC (0.40±0.02) (**Figure 1D**). This difference in NNF may be due to unmeasured venous contributions, such as small lumbar veins, the right GV, or the right suprarenal vein (**Figure 1E**).

### Blood flow in participants with PeVD imaging findings

Blood flow along the abdominopelvic path of venous return was compared between LCIV compression (n=19), GV reflux (n=24), or LRV compression (n=14) participants and NVA participants (n=29) (**Supplemental Figure 2**).

### Disorders involving Iliac Veins

Participants with LCIV compression displayed significantly diminished LCIV NNF compared to participants with NVA (LCIV compression: 0.38±0.03 vs. NVA: 0.47±0.02, p=0.03). LCIV compression appeared to increase the RCIV NNF compared to NVA (LCIV compression: 0.67±0.04 vs. NVA: 0.54±0.02, p=0.01), leading to similar IVC confluence NNF (LCIV compression: 1.0±0.05 vs. NVA: 1.1±0.07, p=0.54) (**Figure 2A**). The LCIV/RCIV flow ratio was significantly decreased in LCIV compression compared to the ratio in NVA participants (LCIV compression: 0.62±0.07 vs. NVA: 0.92±0.05, p=0.001) (**Figure 2B**). Despite asymmetric flow in the CIVs, conservation of flow at IVC confluence was observed in participants with LCIV compression (**Figure 2C**). Compared to NVA, LGV reflux or LRV compression groups showed no significant difference in NNF at the LCIV, RCIV, or IVC confluence (**Figure 2A-B**). Of the 7 LCIV compression participants who also underwent IVG, most had concordant findings with MRI. IVG detected LCIV compression in 2 participants without the morphologic finding on MRI. (**Table I**).

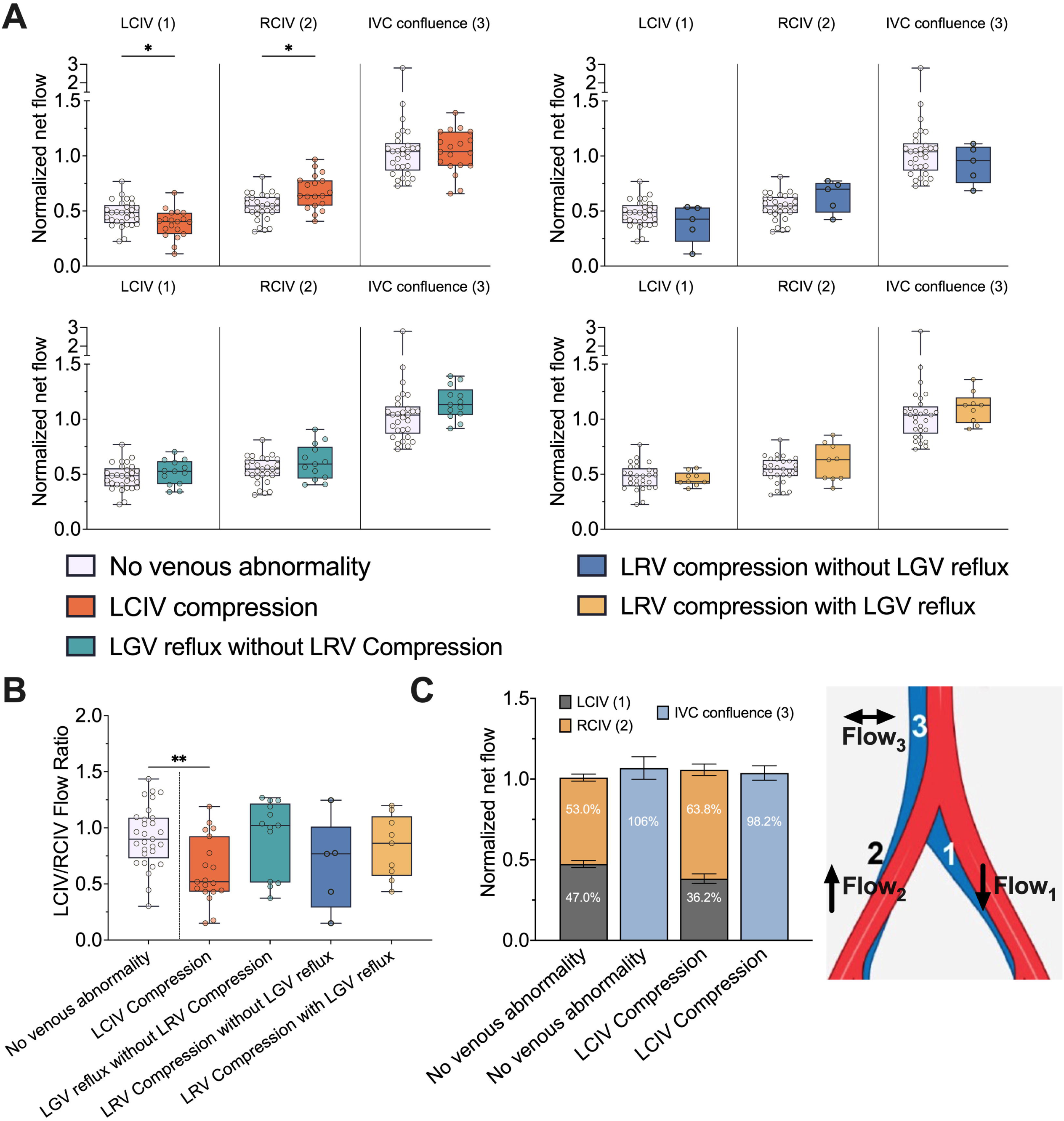

### Disorders involving Renal and Gonadal Veins

In participants with both LRV compression and LGV reflux, NNF was significantly diminished at the juxta-caval LRV (0.13±0.06) compared to NVA (0.45±0.03, p<0.001). Participants with LGV reflux without LRV compression (0.34±0.05, p=0.14 vs. NVA) and LRV compression without LGV reflux (0.39±0.05, p=0.62 vs. NVA) did not have significantly decreased juxta-caval LRV NNF. Among these groups, the NNFs at the juxta-caval RRV or perihilar renal veins were similar (**Figure 3A**). The LRV flow ratio was significantly decreased in LRV compression with LGV reflux compared to NVA (LRV compression: 0.39±0.17 vs. NVA: 1.3±0.13 p<0.001) (**Figure 3B**). No significant difference was observed in NNF in renal veins of participants with LCIV compression compared to NVA (**Figure 3A-B**). Of the 7 participants with LRV compression who also underwent IVG, most had discrepant findings between the modalities. LRV compression was observed on IVG in 1 participant without the morphologic finding on MRI. Five LRV compression participants had the finding on MRI, but not IVG. Of 14 participants with any GV reflux, IVG detected reflux in 5 participants without the morphologic finding on MRI. One participant had the finding on MRI, but not IVG (**Table I**).

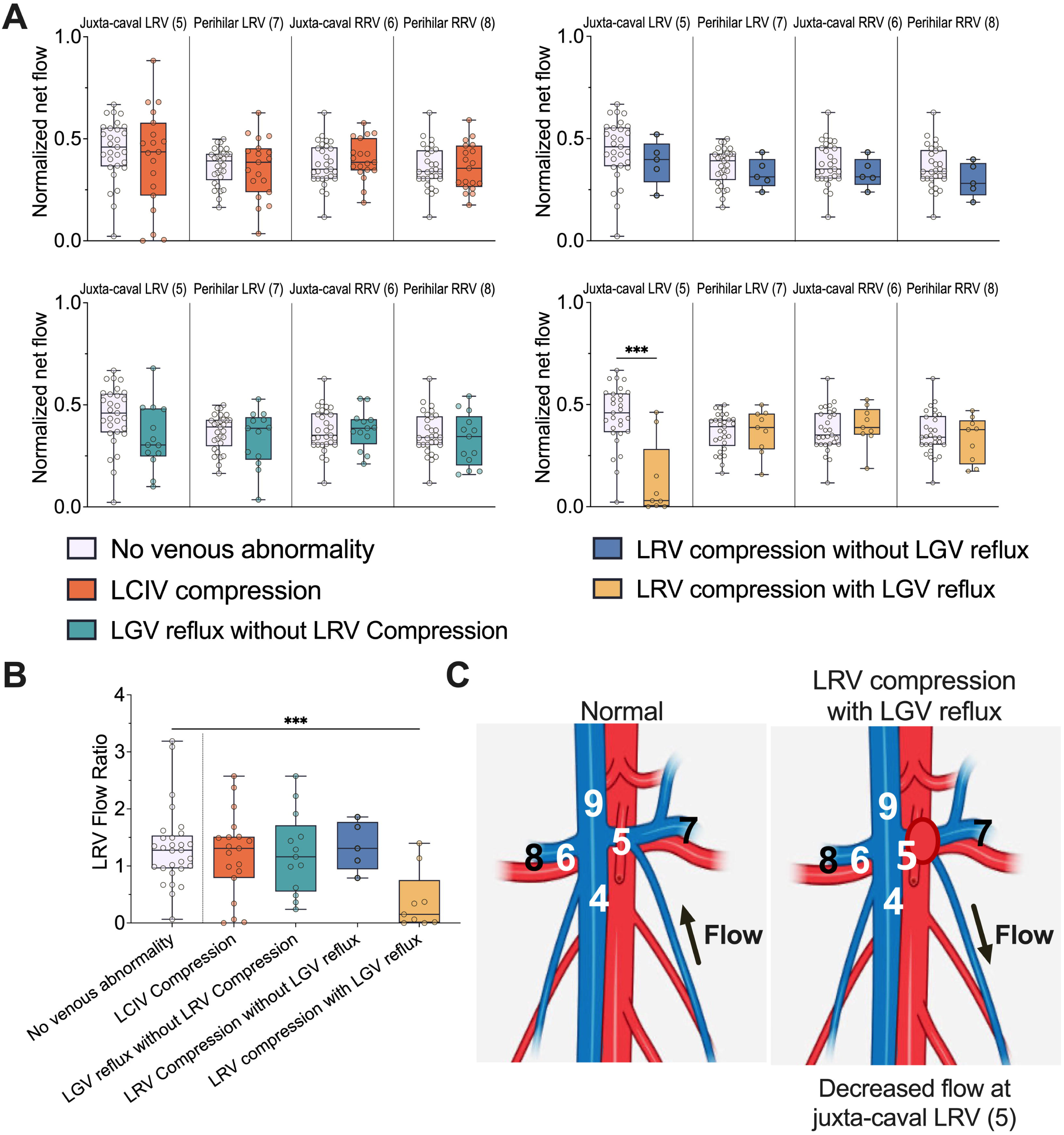

### Alternate routes of blood flow in compressive etiologies of PeVD

Participants with LCIV compression were subclassified by the presence (n=11) or absence (n=8) of pelvic venous collaterals. The LCIV NNF decreased in participants with LCIV compression with collaterals (0.32±0.03) compared to those without collaterals (0.47±0.04) and those with NVA (0.47±0.02), with significant decrease compared to NVA (p=0.003). The RCIV NNF also was increased in participants with LCIV compression with collaterals (0.72±0.05) compared to those without collaterals (0.62±0.04) and those with NVA (0.54±0.02), and significant increase in RCIV NNF compared to that in NVA (p=0.01). The asymmetry of NNF between the LCIV and RCIV was greatest in participants with LCIV compression with collaterals (p=0.006) (**Figure 4B-C**).

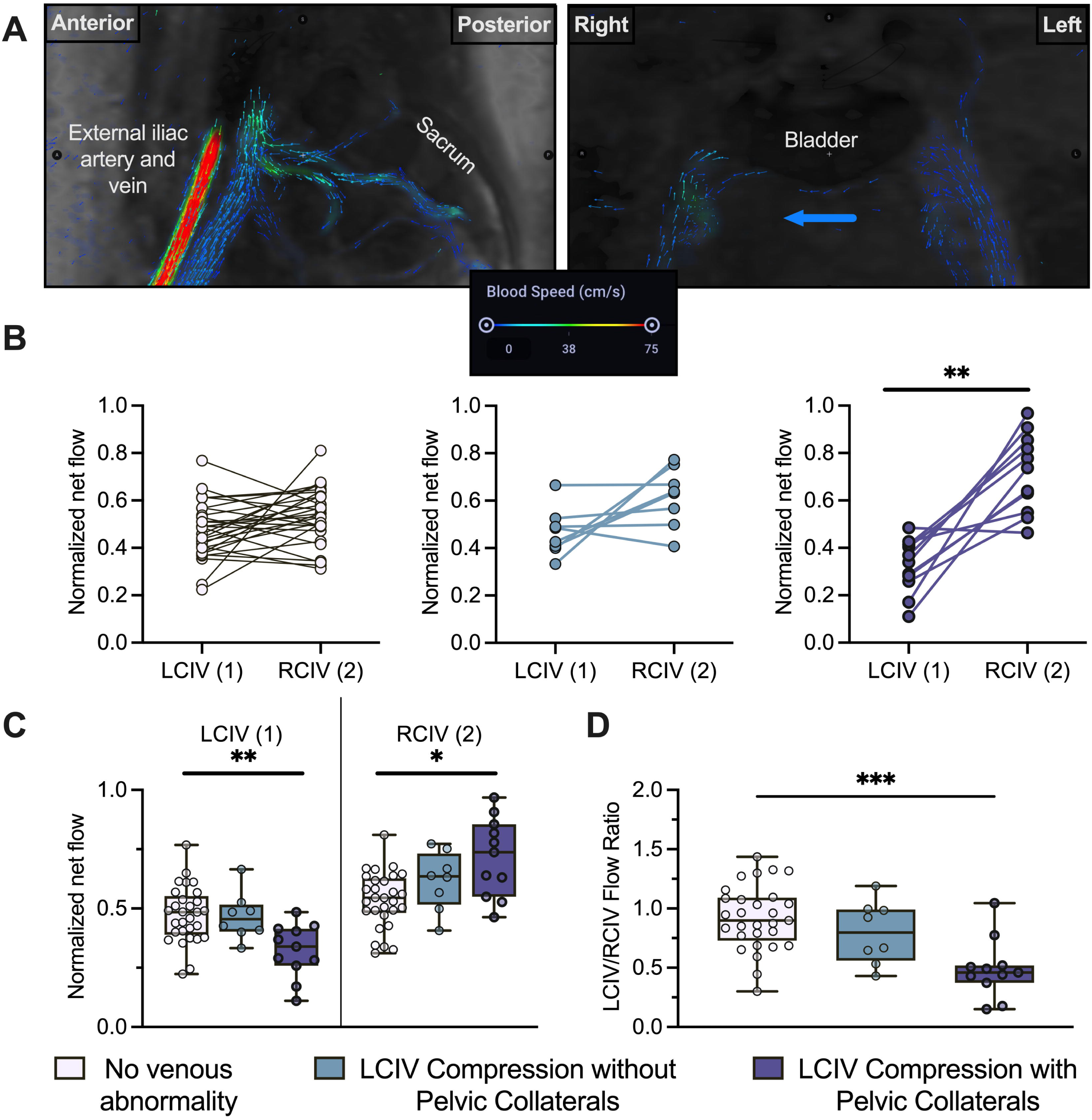

The LCIV/RCIV flow ratio (with pelvic collaterals: 0.49±0.08, without pelvic collaterals: 0.80±0.09) was significantly decreased in LCIV compression with pelvic collaterals compared to NVA (p=0.0005). The LCIV/RCIV flow ratio in LCIV compression without pelvic collaterals was intermediate between the NVA and the LCIV compression with pelvic collaterals groups (p>0.1 compared to NVA and to the “with pelvic collaterals” groups) (**Figure 4D**). The asymmetry of CIV flow in participants with LCIV compression was exacerbated in the presence of alternate paths of flow, pelvic reservoir collaterals.

Participants with LRV compression were stratified by the presence (n=9) or absence (n=5) of LGV reflux. The juxta-caval LRV NF was significantly decreased in participants with LRV compression with LGV reflux (0.13±0.06) compared to those without LGV reflux (0.39±0.05, p<0.01) and those with NVA (0.44±0.03, p=0.0008). Perihilar LRV NNF was similar (p>0.9) in all three groups (LRV compression with LGV reflux: 0.37±0.04, LRV compression without LGV reflux: 0.33±0.03, NVA: 0.36±0.02). Among participants with LRV compression and LGV reflux, the juxta-caval LRV NNF was significantly reduced compared to their perihilar LRV NNF (p=0.04), as opposed to the significantly increased flow at the juxta-caval LRV compared to the perihilar LRV in NVA (p=0.04) (**Figure 5B-C**). The LRV flow ratio was significantly decreased in LRV compression with LGV reflux (0.39±0.17, p=0.004) compared to NVA and compared to LRV compression without LGV reflux (1.3±0.19, p=0.03) (**Figure 5D**).

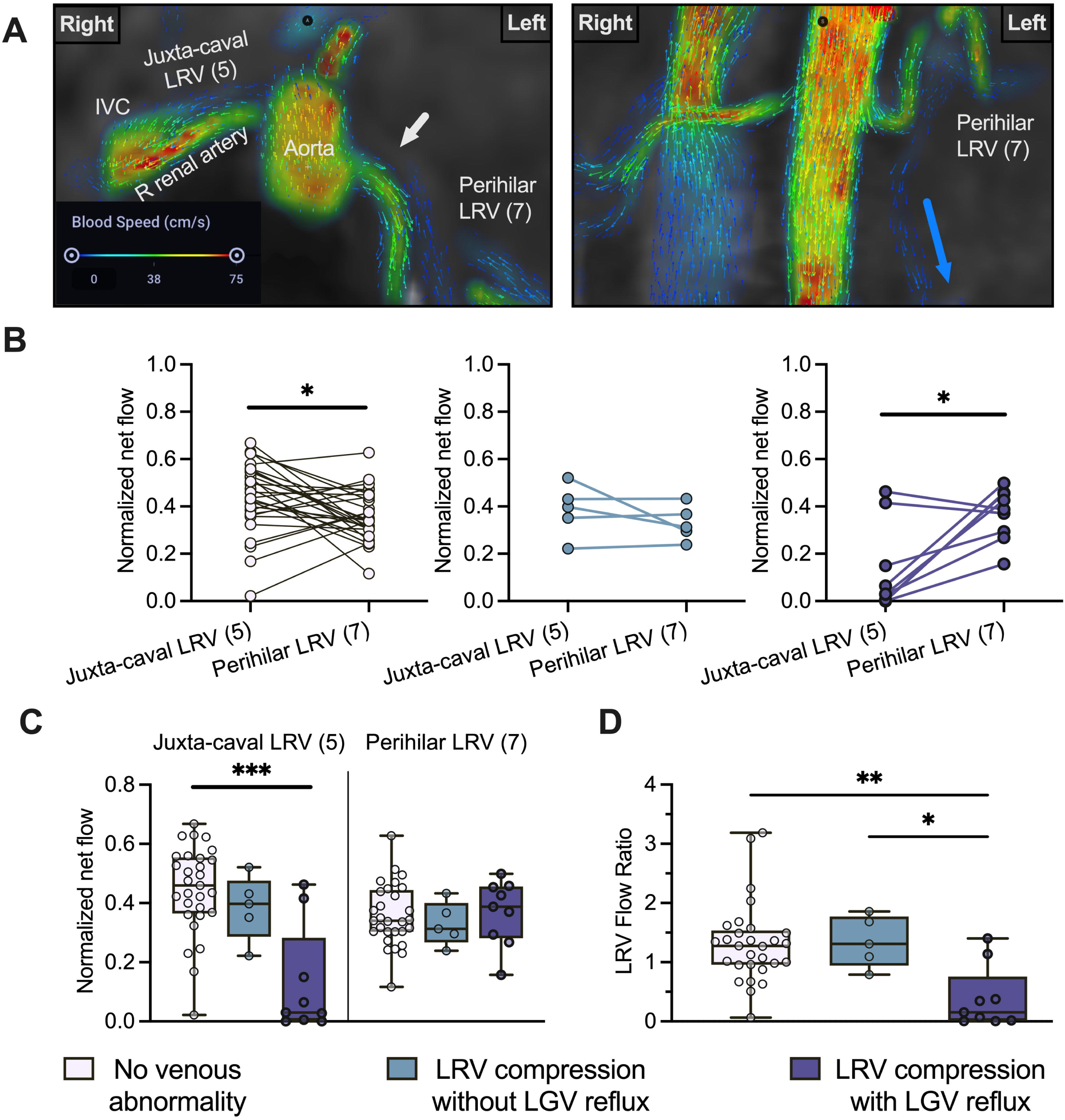

## Discussion and Conclusions

We aimed to determine whether quantitative 4D-Flow MRI can localize and quantify venous blood flow abnormalities in PeVD. The venous flow and speed in NVA participants were concordant with those reported in the literature [13,38–40]. Conserved flow at the IVC and renal venous confluences further validated our methodology [33]. Increased RCIV flow, decreased LCIV flow, and conserved IVC flow in participants with LCIV compression, indicated left-to-right CIV shunting. Asymmetrical flow between the LCIV and RCIV was most pronounced in LCIV compression with visible pelvic collaterals; suggesting exacerbated left-to-right shunting in the presence of pelvic reservoirs—alternate paths of flow. The weaker pattern in LCIV compression without visible pelvic collaterals could indicate less severe compression or flow diversion through other reservoirs, such as lower extremity veins, outside the field-of-view [4,34]. Decreased juxta-caval LRV flow was seen in LRV compression with LGV reflux. Juxta-caval and perihilar LRV flow in LRV compression without LGV reflux was indistinguishable from that of NVA. Reduced juxta-caval LRV flow was not related to decreased left kidney venous outflow, but rather an abnormality in flow along the course of the LRV, such as compression and/or LGV reflux.

As a single-institution retrospective analysis with a limited number of participants, there were important limitations. The NVA group were not true healthy controls. Wider variability in quantitative data for PeVD relative to the NVA increases the difficulty of detecting differences. VENC selection can affect flow and speed measurements; however, reliable flow and velocity measures in veins have been obtained at similar VENC (∼100 cm/s) [14,39]. Flow measurements were not correlated with another modality; nonetheless, previous work has demonstrated strong concordance between 4D-Flow MRI and IVG, ultrasound, and 2D phase-contrast MRI [41–45]. Comparison of IVG and 4D-Flow MRI was limited, with 17 participants undergoing both. IVUS was variably used in the IVG procedures, another important limitation given the high resolution in assessing venous occlusion. Discrepant detection of LRV compression and GV reflux could reflect GV reflux exacerbation by more upright positioning—available on IVG, but not on MRI. LRV compression by soft tissue structures may be more salient on cross-sectional imaging than IVG.

Quantitative 4D-Flow MRI measurements reflected morphological imaging findings, distinguishing among normal and PeVD venous flow patterns, identifying potential treatment targets. The relationship between venous flow abnormalities and clinical symptoms remains unclear, but it is a critical future research direction. Nonetheless, 4D-Flow MRI provides large field-of-view 3D quantitative data. These potential biomarkers could facilitate disease severity classification, recurrence monitoring, and triaging patients for IVG. IVG often coupled with IVUS, a more focused and invasive exam, and 4D-Flow MRI could be complementary together.

## Data Availability

All data produced in the present study are available upon reasonable request to the authors.

## Acknowledgements

Farhoud Faraji, MD, PhD for assistance with visual abstract graphic design

## Funding Declaration

This study was funded in part by NIH grant T32EB00597.

## Manuscript Type

Clinical Investigation

## List of abbreviations

PeVD: Pelvic venous disorder
NVA: no vascular abnormalities
LCIV: left common iliac vein
GV: gonadal vein
LGV: left gonadal vein
LRV: left renal vein
IVG: invasive venography
POTS: postural orthostatic tachycardia syndrome
MTS: May-Thurner syndrome
4D-Flow MRI: three-dimensional time-resolved phase-contrast MRI with velocity encoding
NNF: normalized net flow

## Compliance with Ethical Standards

Conflict of interest statement: Disclosures of conflicts of interest: C.E.B. disclosed no relevant relationships. S.W. disclosed no relevant relationships. P.R.T. consulting fees from Amgen, Novo-Nordisk, Medtronic, Boehringer-Ingelheim, Sanofi, and Esperion Therapeutics; shareholder in Epirium Bio. A.C.R. disclosed no relevant relationships. C.I. disclosed no relevant relationships. A.H. grants from GE Healthcare and Bayer unrelated to the present work; patent filed by Stanford University and licensed by multiple vendors related to phase-error correction and analysis of 4D flow MRI; cofounder of and shareholder in Arterys. J.L. disclosed no relevant relationships.

For this type of study formal consent is not required. This study has obtained IRB approval from (UCSD IRB: #201979) and the need for informed consent was waived.

For this type of study consent for publication is not required.

## Notes

### Author Declarations

This study has obtained IRB approval from (University of California, San Diego IRB: #201979).

### Summary of Updates

Updates to the introduction and discussion to include consideration of intravascular ultrasound

