## Supplemental Methods for "Quantitative 4D-Flow MR Imaging of Abdominopelvic Vasculature in Pelvic Venous Disorders"

### Imaging segmentation

Juxta-caval renal vein measurements were acquired on the renal vein just before it joined the IVC. Perihilar RV measurements were obtained at the renal vein just medial to the hilum. On the LRV, the left GV (LGV) is expected to join with the LRV between the perihilar LRV and the juxta-caval LRV. Proximal LCIV measurements were acquired on the LCIV just inferior to the cross-over of the common iliac artery. RCIV measurements were obtained at approximately the same level as the LCIV measurements. Suprarenal aorta measurements were taken superior to the origin of the celiac axis (**Figure 1A**).

*Blood flow in participants with participants without venous abnormality and PeVD imaging findings*

Abdominopelvic non-normalized net flow (mL/s), maximum peak speed (cm/s), and NNF at the suprarenal IVC, infrarenal IVC, juxta-caval LRV and RRV, perihilar LRV and RRV, IVC confluence, LCIV, and RCIV in participants without venous abnormality (NVA) were evaluated. To obtain NNF, the mean net venous flow at each location was normalized to the mean net flow at suprarenal aorta per participant. To assess potential effects on measurements based on VENC, NVA participants were categorized based on VENC level and the mean net flow, maximum peak speed, and NNF were quantified for each group. NNFs were also evaluated in groups having GV reflux, LCIV compression, or LRV compression.

*Conservation of flow at venous confluences in participants without venous abnormality*

To assess conservation of flow in participants with NVA, NNF at venous confluences was compared to the summation of the NNF measured at the contributing veins. Percentages of the summed contributing vein NNF were calculated for each contributing vein and the venous confluence. The IVC confluence NNF was compared to the sum of the RCIV NNF and LCIV NNF. The suprarenal IVC NNF was compared to the sum of the infrarenal IVC, juxta-caval LRV, and juxta-caval RRV NNFs.

*Disorders involving Iliac Veins*

The IVC confluence, LCIV, and RCIV NNFs were evaluated in participant group with morphologic findings of NVA, LCIV compression, LGV reflux without LRV compression, LRV compression without LGV reflux, or LRV compression with LGV reflux. The NNF per participant was calculated by dividing the mean net LCIV, RCIV, IVC confluence flows by mean net flow at the aortic bifurcation. The LCIV/RCIV flow ratio is the NNF at the LCIV divided by NNF at the RCIV. The conservation of flow assessment was performed as described previously for the NVA and LCIV compression groups. To assess the effects of alternate routes of flow on iliac venous flow, the RCIV and LCIV NNFs and LCIV/RCIV flow ratio in participants with NVA, LCIV compression without pelvic collaterals, or LCIV compression with pelvic collaterals were examined.

*Disorders involving Renal and Gonadal Veins*

The juxta-caval LRV and RRV and perihilar LRV and RRV NNFs were evaluated in participant groups with morphologic findings of NVA, LCIV compression, LGV reflux without LRV compression, LRV compression without LGV reflux, or LRV compression with LGV reflux. The renal venous flow was normalized to the summed mean net flow of the bilateral renal arteries to obtain the NNF per participant. The LRV flow ratio was the NNF at the juxta-caval LRV divided by NNF at the perihilar LRV. To assess the effects of LGV reflux, as an alternate route of blood flow, on LRV flow, the juxta-caval and perihilar LRV NNFs and LRV flow ratio in participants with NVA, LRV compression without LGV reflux, or LRV compression with LGV reflux were examined.
