## Supplemental Figures 1 and 2 for "Quantitative 4D-Flow MR Imaging of Abdominopelvic Vasculature in Pelvic Venous Disorders"

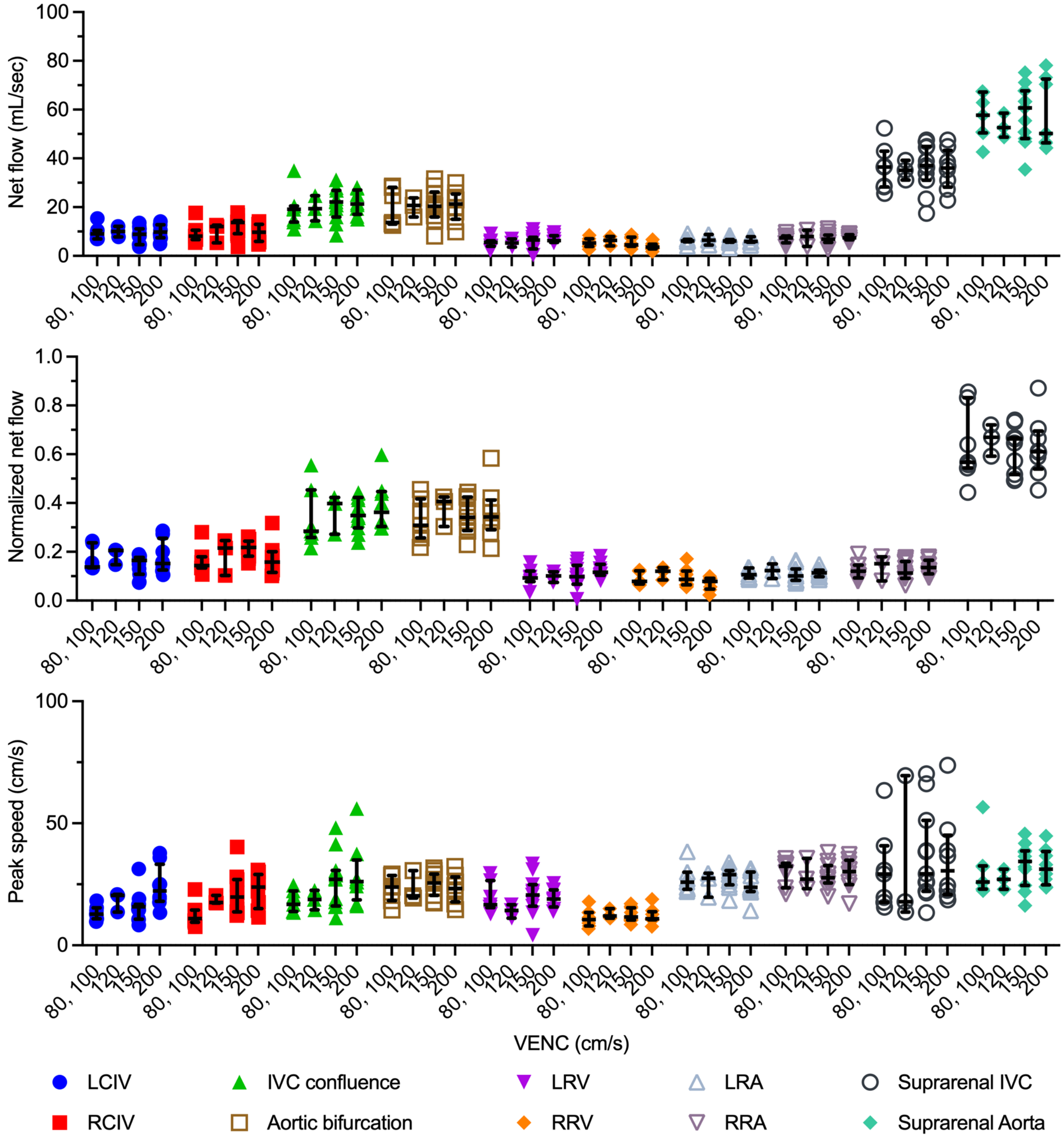


**Supplemental Figure 1. Flow and speed in NVA participants stratified by VENC setting.** Non-normalized net flow, NNF (normalized to suprarenal aorta net flow), and peak speed in NVA participants (n = 29).Participants grouped by VENC setting of their 4D-Flow MRI exam (80 cm/s: n=1, 100 cm/s: n=6, 120 cm/s: n=3, 150 cm/s: n=11, or 200 cm/s: n=8). Lines depict median (middle line), interquartiles (25th and 75th percentiles, ends of the lines). Each dot represents a participant. IVC, inferior vena cava; LCIV, left common iliac vein; RCIV, right common iliac vein; LRV, left renal vein; RRV, right renal vein; LRA, left renal artery; RRA, right renal artery; NVA, no venous abnormality; NNF, normalized net flow; VENC, velocity encoding.

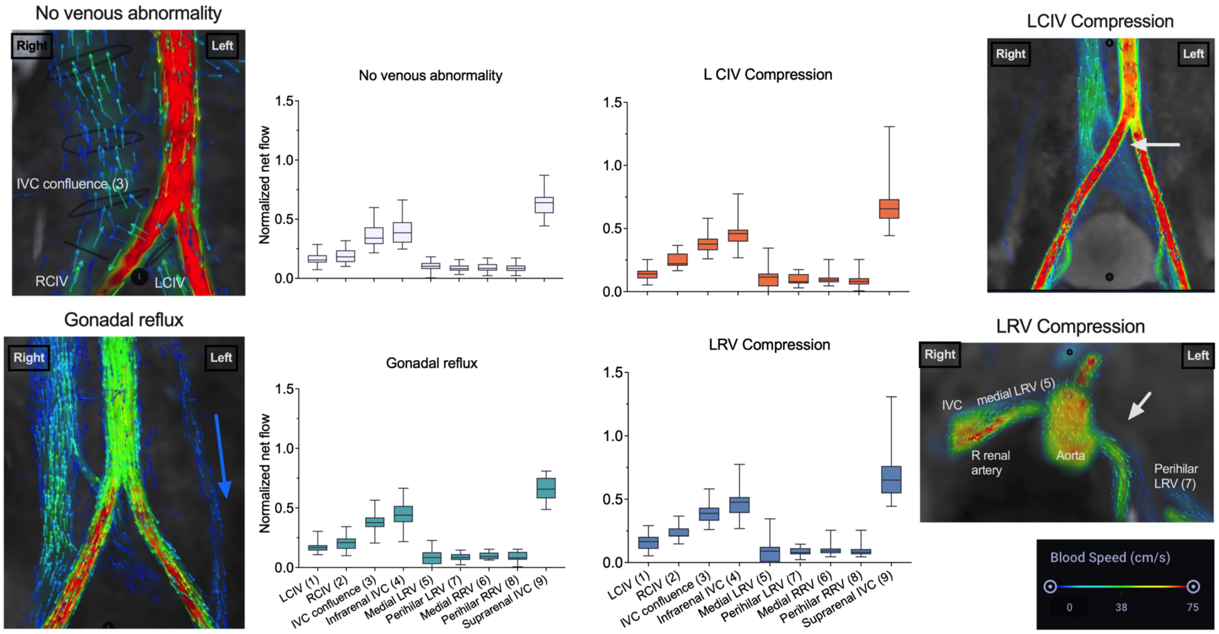


**Supplemental Figure 2. Abdominopelvic flow in participants with PeVDs.** *Left upper*, Coronal 4D-Flow MR image with vectors and color-coded speed from NVA participant. Graph depicting NNF in NVA participants (n = 29). *Right upper*, Coronal 4D-Flow MR image from participant with LCIV compression (arrow). Graph depicting NNF (n = 19). *Left lower,* Coronal 4D-Flow MR image from participant with LGV reflux (arrow in direction of blood flow). Graph depicting NNF (n = 24). *Right lower,* Axial 4D-Flow MR image from participant with LRV compression (arrow). Graph depicting NNF (n = 14). Speed (cm/s) color scale for all 4D-Flow images. For the plots of abdominopelvic venous flow from iliac veins to the suprarenal IVC, NNF for each vessel position was computed by dividing the mean net venous flow by the mean net suprarenal aorta flow in each subject was used for normalization to account for individual differences in baseline vascular states. All box and whisker plots depict the median (middle line), interquartiles (25th and 75th percentiles, ends of the boxes) and range (whisker ends). CIV, common iliac vein; IVC, inferior vena cava; LCIV, left common iliac vein; PeVD, pelvic venous disorder; RCIV, right common iliac vein; LRV, left renal vein; RRV, right renal vein; NVA, no venous abnormality; NNF, normalized net flow.
